# The Hidden Architecture of Brain Structural Variability in 22q11.2 Deletion Syndrome: A Multi-site Study

**DOI:** 10.64898/2026.05.18.26353539

**Authors:** Rune Boen, Kathleen P. O’Hora, Hoki Fung, Leila Kushan, Charles H. Schleifer, Tyler E. Dietterich, Carolyn M. Amir, Samuel Klein, Jee Won Kang, Haley R. Wang, Dylan E. Hughes, Julio E. Villalon-Reina, Melody J.Y. Kang, Yanghee Im, Kuldeep Kumar, Dag Alnæs, Kathleen Angkustsiri, Kevin M. Antshel, Geor Bakker, Anne S. Bassett, Nancy J. Butcher, Linda E. Campbell, Samuel J.R.A. Chawner, Eva W.C. Chow, Michael C. Craig, Nicolas A. Crossley, Eileen Daly, Fabio Di Fabio, Joanne L. Doherty, Beverly S. Emanuel, Ania M. Fiksinski, Jennifer K. Forsyth, Marianna Frascarelli, Wanda P. Fremont, Maria Gudbrandsen, Raquel E. Gur, Joachim F. Hallmayer, Maria Jalbrzikowski, Wendy R. Kates, David E. Linden, Kathryn L. McCabe, Donna M. McDonald-McGinn, Declan Murphy, Kieran C. Murphy, Ruth O’Hara, Michael J. Owen, Allan L. Reiss, Gabriela M. Repetto, David R. Roalf, Kosha Ruparel, J Eric Schmitt, Sam A. Sievertsen, Tony J. Simon, Zachary H. Trevorrow, Therese van Amelsvoort, Marianne B.M. van den Bree, Jacob A.S. Vorstman, Elaine H. Zackai, Christopher R.K. Ching, Paul M. Thompson, Carrie E. Bearden, the ENIGMA-22q Working Group

**Author notes:** Correspondence: Carrie E. Bearden, A7-460 Semel Institute, Los Angeles, CA 90095, USA.

## Abstract

**Importance:** 22q11.2 deletion syndrome (22q11DS) is among the strongest genetic risk factors for neuropsychiatric disorders and has marked effects on brain structure. Yet, it remains unclear which neuroanatomical features reflect uniform effects of the deletion versus inter-individual biological processes relevant to psychiatric outcomes. Identifying these features is critical for developing targeted treatments and interventions.

**Objective:** To identify brain regions where 22q11DS exerts its most consistent and most variable impacts, and to test whether these patterns align with normative neurotransmitter receptor distributions and cortical growth trajectories.

**Design:** Multisite cross-sectional case-control study.

**Setting:** T1-weighted brain MRI data were obtained across 15 scanners. MRI data underwent standardized processing, quality control procedures and statistical site-adjustment using ComBat.

**Participants:** A total of N = 438 individuals with 22q11DS (5-54 years, 48% females) and 380 typically developing controls (6-58 years, 48% females).

**Main Outcomes and Measures:** Primary outcomes were global and regional cortical thickness and surface area. Mean and dispersion estimates were calculated using double generalized linear models, correcting for age, age^2^, sex (and intracranial volume for surface area). Quantile shift functions characterized fine-scale distributional differences. Sensitivity analyses adjustedt for co-occurring neuropsychiatric disorders, antipsychotic use and deletion subtype. Secondary outcomes included spatial correspondence between regional structural alterations and normative maps of neurotransmitter receptor density and cortical expansion.

**Results:** Compared with controls, individuals with 22q11DS showed widespread mean differences in cortical thickness and surface area. Notably, 22q11DS was associated with greater regional heterogeneity in both measures, except for reduced dispersion in the anterior cingulate. Effects were attenuated after covariate adjustment. Cortical thickness differences spatially overlapped with regions enriched for glutamatergic and GABAergic receptors. There was partial evidence linking surface area dispersion patterns to normative cortical growth trajectories.

**Conclusions and Relevance:** 22q11DS exerts broad effects on cortical structure consistent with a global developmental mechanism, reflected in widespread mean shifts. Beyond these, region-specific variability, particularly in cortical thickness, suggests individualized neurobiological processes. The anterior cingulate emerges as a region of consistent structural deviation. Overall, structural variability in 22q11DS aligns with normative patterns of excitatory-inhibitory signaling and cortical development, implicating these pathways as potential targets for intervention.

**Key points:** *Question:* Is 22q11.2 deletion syndrome (22q11DS) associated with altered spatial heterogeneity of cortical structure, and do these patterns map onto the brain’s underlying neurochemical and developmental architecture?

*Findings:* In this multisite case-control study of 438 22q11DS and 380 controls, 22q11DS showed regionally patterned increases in cortical thickness heterogeneity and lower cortical surface area heterogeneity. Spatial patterns of cortical thickness differences were aligned with cortical gradients of glutamate and GABAergic receptor density.

*Meaning:* Although 22q11DS is generally associated with reduced brain volume and increased cortical thickness, the present findings reveal regionally patterned cortical alterations aligned with neurotransmitter-specific cortical organization, suggesting a mechanistic link between excitatory–inhibitory signaling architecture and individualized neuroanatomic effects of 22q11DS.

## Introduction

22q11.2 deletion syndrome (22q11DS) is among the most common human microdeletions, affecting approximately 1 in 2,000-4,000 live births (1,2). 22q11DS presents with striking clinical heterogeneity, encompassing cardiac malformations, developmental delays, and markedly increased risk for neurodevelopmental disorders, notably psychotic and autism spectrum disorders (1,3–5). Neuroimaging studies have consistently identified a structural brain signature in 22q11DS, characterized by thicker cortex and smaller cortical surface area compared to controls (6). However, the wide variability in clinical outcomes suggests that group-level mean differences alone may not fully capture the neurobiological effects of the deletion (1,3,5,7,8). Emerging evidence across developmental neuropsychiatric disorders suggests that affected individuals often show increased inter-individual variability in brain structure (9,10). Thus, modeling both the mean and the dispersion of neuroimaging features may provide additional insight into the mechanisms underlying variable clinical expression in 22q11DS.

Cortical thickness (CT) and surface area (SA) reflect distinct and partially independent neurodevelopmental processes (11). SA is largely established early in life via progenitor cell proliferation (12,13). In contrast, CT is more dynamic across development and reflects a composite of processes including dendritic arborization, synaptic density and pruning, and myelination(14,15). The widespread cortical thickening in 22q11DS (6) may therefore reflect disruptions in later-emerging processes related to circuit refinement (16). Converging evidence from cellular and animal models implicates alterations in excitation–inhibition (E/I) balance - especially involving GABAergic and glutamatergic signaling - in shaping these processes and influencing cortical maturation (17–20). Accordingly, variation in CT in 22q11DS may, in part, reflect alterations in E/I-regulated mechanisms (21–23), which are also implicated in psychosis and autism spectrum disorders (24–29).

In contrast, reductions in SA are approximately twice the magnitude of thickness effects (6) and may reflect early developmental constraints that produce more uniform alterations across individuals. At the same time, regional variability in SA may depend on the developmental trajectory of cortical expansion. Association cortices, particularly frontotemporal regions, undergo prolonged development and exhibit disproportionate (hyperallometric) expansion, whereas primary sensory and limbic regions follow more constrained developmental trajectories (30–32). These differences suggest that genetic disruptions may produce greater variability in regions with extended developmental windows, and more consistent effects in regions with more limited expansion.

Despite these distinctions, most neuroimaging studies of 22q11DS and other rare variants have focused on mean differences, leaving variability largely unexplored. Characterizing both central tendency and dispersion may be particularly informative in genetically defined conditions, where a shared genetic alteration leads to heterogeneous outcomes. Identifying brain regions that show consistent versus variable effects may help distinguish core features of the syndrome from those reflecting downstream or individual-specific processes. Here, we extend prior work from the ENIGMA-22q consortium(6) by incorporating additional cohorts and applying double generalized linear models to simultaneously quantify both the mean and the dispersion of these cortical features. We hypothesized that cortical thickness alterations would preferentially localize to regions enriched for glutamatergic and GABAergic receptor density, consistent with an excitation–inhibition imbalance framework. Conversely, we expected cortical surface area alterations to follow normative scaling principles of cortical expansion (30), showing spatial overlap with evolutionary, developmental, and allometric gradients of cortical growth.

## Methods

### Design and Participants

We conducted a multisite case-control magnetic resonance imaging (MRI) study on global and regional cortical thickness and cortical surface area measures in 22q11DS. A total of 818 participants (22q11DS = 438, Controls = 380) across 15 scanner sites were included in the study (**Table 1**), after exclusion of subjects from sites that had no control participants (see eMethods for details). All participants gave informed consent (and/or assent from minors), and sites obtained approval from local ethics committees.

**Table 1.** Sample demographics.

|  |  | Group |  | P-value |
| --- | --- | --- | --- | --- |
|  |  | Control<br>(N = 380) | 22q11DS<br>(N = 438) |  |
| Age (y) | Min / Max | 6.0 / 58.0 | 5.0 / 54.0 | 0.096 |
|  | Med [IQR] | 18.0 [12.7;23.0] | 17.0 [12.0;22.0] |  |
|  | Mean (std) | 19.5 (9.8) | 18.3 (8.9) |  |
| Sex (%) | Female | 182 (48%) | 211 (48%) | 0.937 |
|  | Male | 198 (52%) | 227 (52%) |  |
| Site | Cardiff | 11 | 15 | - |
|  | IoP | 25 | 37 |  |
|  | Newcastle | 23 | 16 |  |
|  | Maastricht | 45 | 27 |  |
|  | Penn | 48 | 49 |  |
|  | Rome | 26 | 17 |  |
|  | Santiago | 28 | 25 |  |
|  | Stanford | 6 | 6 |  |
|  | SUNY | 20 | 54 |  |
|  | Toronto1 | 12 | 15 |  |
|  | UCDavis1 | 38 | 40 |  |
|  | UCDavis2 | 48 | 64 |  |
|  | UCLA_BMC | 23 | 28 |  |
|  | UCLA_CCN | 19 | 34 |  |
|  | UCLA_PRISMA | 8 | 11 |  |
| ASD | ASD- | 380 (100%) | 78 (50%) | - |
|  | ASD+ | 0 (0%) | 78 (50%) |  |
| Psychotic Disorder | Psy- | 380 (100%) | 386 (88%) | - |
|  | Psy+ | 0 (0%) | 52 (12%) |  |
| Deletion type | Typical (A-D) | - | 249 (88%) |  |
|  | Atypical | - | 34 (12%) |  |

*Note.* Autism spectrum disorder (ASD+) and psychotic disorder (PSY+) diagnosis indicate individuals with a confirmed diagnosis for whom diagnostic information was available. 282 22q11DS participants in the current analysis were not formally assessed for ASD. 155 22q11DS participants had missing data for deletion size. Atypical = any smaller 22q11.2 deletion variant than the typical A-D deletion.

### MRI Processing

3D volumetric T1-weighted brain images were obtained for 22q11DS and controls across sites, and preprocessed following the ENIGMA neuroimaging pipeline (https://github.com/ENIGMA-git) to derive global and 68 regional measures of cortical thickness and cortical surface area using the Desikan-Killiany atlas. The processed data underwent cortical quality control (QC) following the ENIGMA QC pipeline (https://enigma.ini.usc.edu/protocols/imaging-protocols/) and was statistically adjusted for site-related variation using ComBat (33) (eMethods). After QC, 401 22q11DS and 351 control participants were retained for analyses.

### Statistical Analysis

All statistical analyses and data visualization were performed in R version 4.4.1 or MATLAB (R2024B, The MathWorks, Inc., Natick, Massachusetts, United States). We used double generalized linear models to estimate group differences in both the mean and dispersion of cortical measures. For each feature, we fitted a double generalized model with group status (22q11DS vs controls) as the factor of interest and age, age^2^, sex and intracranial volume (ICV; only for SA measures) as covariates. This allows the mean and variance of the cortical measures to be modeled simultaneously in a two-part regression framework (9,34). Of note, the dispersion model tested whether the variance of CT or SA differ between groups. The *p*-values underwent false discovery rate (FDR) correction across all group comparisons per modality. All cortical features were Z-transformed using the mean and standard deviation derived from the control group. Furthermore, we visualized the differences in quantiles between 22q11DS and controls using the *rogme* package (https://github.com/GRousselet/rogme) by utilizing the shift function with bootstrapping (n = 1000), including confidence intervals for the quantile differences. For visualization of the results, the standardized mean differences for significant regions were mapped using the *ggseg* package (35). All primary analyses were performed QCed data (see sFigure1 and eTable 1 for pre-QC results).

### Robustness analyses

Variability in neuroimaging features can be influenced by image quality (36), neuropsychiatric diagnoses(including autism(37) and psychosis spectrum disorders(38)), and medication (39). Further, prior work from the ENIGMA-22q consortium has revealed differences in cortical structure as a function of deletion subtype(6). In secondary analyses, we adjusted for diagnoses of psychosis and autism spectrum disorders, atypical deletion (i.e., smaller than the typical A-D deletion), and reported use of antipsychotic medication (i.e., all variables were binary coded). These analyses were performed to account for any systematic mean shifts that could be due to secondary disorder-related mechanisms, while preserving within-group variability. Analyses that included diagnostic status and antipsychotic medication were performed separately due to high collinearity between these variables.

### Spatial overlap of cortical thickness with glutamate/GABAergic maps

To investigate neurobiological mechanisms underlying differences in both the mean and dispersion of cortical morphology in 22q11DS, we used spin permutation testing, implemented in the ENIGMA Toolbox (40), to quantify the spatial overlap between mean and dispersion cortical maps and cortical gradients from Neuromaps (41,42). Given the hypothesized excitation-inhibition imbalance in 22q11DS and neuropsychiatric disorders (24–29), we focused on glutamate and GABAergic receptor density maps in relation to CT maps. These maps represent normative PET values derived from Neuromaps, including the two glutamate (N-methyl-D-aspartate (NMDA) and metabotropic glutamate receptor subtype 5 (mGluR5) and the one GABA (GABA_A_) receptor density map that were available (42), to index excitatory and inhibitory neurotransmitter systems.

### Spatial overlap of cortical surface area with developmental expansion maps

To probe the developmental origins of altered SA in 22q11DS, we examined spatial correspondence with maps of evolutionary and developmental expansion, as well as allometric scaling, reflecting ontogenetic and phylogenetic cortical expansion patterns (eMethods). The allometric scaling maps consisted of two maps derived from two independent typically developing youth cohorts, i.e., cross-sectional data from the Philadelphia Neurodevelopmental Cohort (PNC) and longitudinal data from the National Institutes of Health (NIH) samples (30,41), which represent how regional brain measures scale in relation to global brain size.

## Results

The results from the double generalized linear models confirmed the previous robust global CT and SA differences between 22q11DS and controls. On average, individuals with 22q11DS had a significantly thicker cortex, lower SA and lower intracranial volume compared to controls, but did not differ in within-group variance (sFigure 2A-D).

### Regional mean and dispersion differences in 22q11DS compared to controls

We observed higher CT in most regions and focal thinning in the superior temporal gyrus, posterior cingulate, and parahippocampal gyrus in 22q11DS compared to controls, consistent with our previous results (6). In addition, the 22q11DS group showed significantly greater within-group variability in thickness of the bilateral entorhinal cortex, rostral anterior cingulate cortex, and left precentral gyrus compared to controls (Figure 1A).

**Figure 1.**
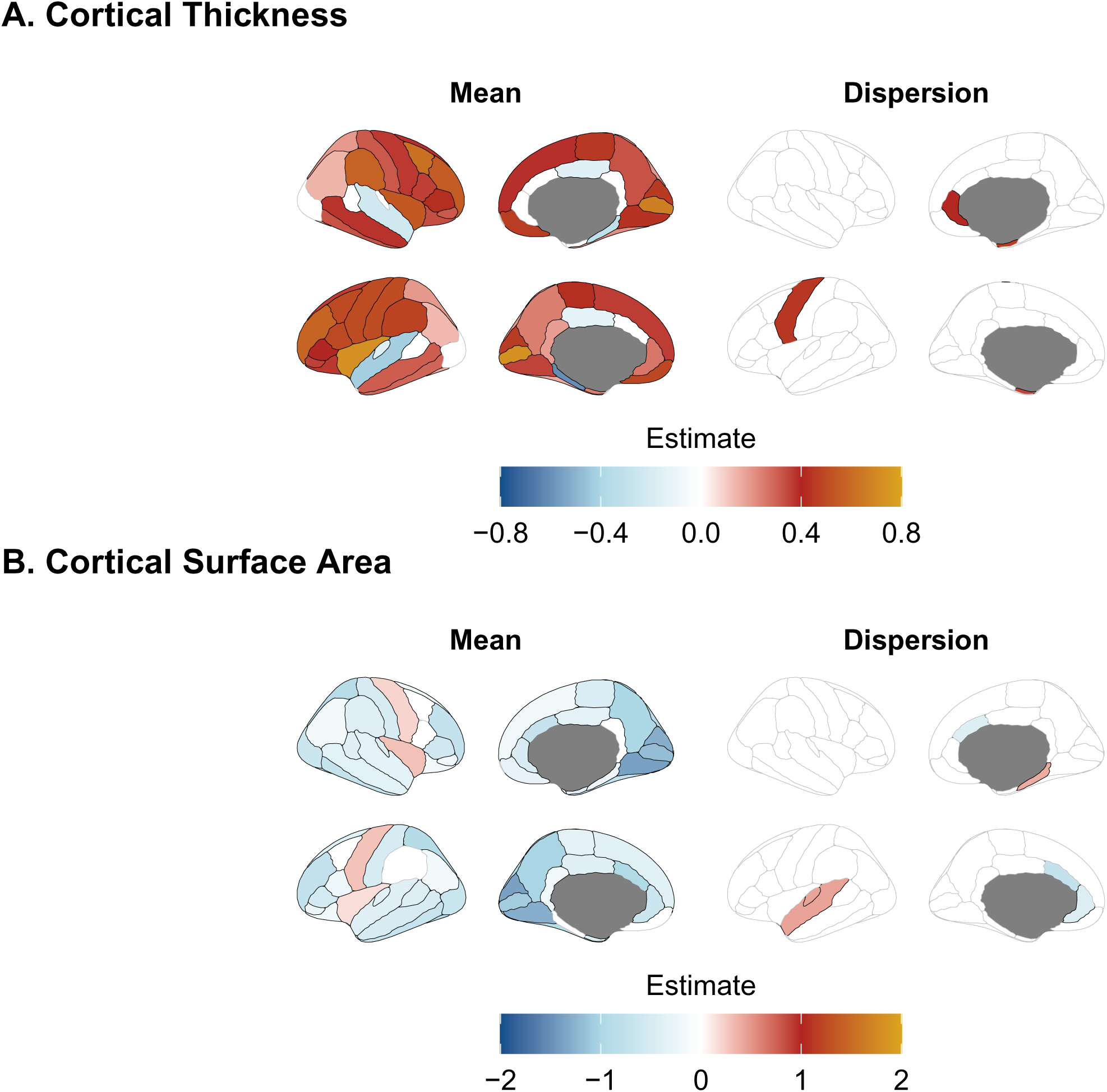
Regional mean and dispersion effects. Regional mean and dispersion effects for A. cortical thickness and B. cortical surface area, adjusted for age, age^2^ and sex (+ intracranial volume for cortical surface area) comparing 22q11DS to controls. The estimate reflects the z-standardized difference between the two groups for the regions that survived FDR-correction, with the control group as the reference. The z-transformation is based on the mean and standard deviation derived from the control group for each regional feature. Mean map: colored regions represent significant group differences in mean values, indicating that 22q11DS group differs on average from controls. Dispersion map: colored regions represent significant group differences in dispersion values, indicating that 22q11DS differs in terms of their variability around the mean from the controls. Positive (red-orange) estimates indicate higher values, and negative (blue) estimates indicate lower values, respectively, in individuals with 22q11DS compared to controls. The overall model estimates are reported in eTable 2.

As expected (6), we also found widespread lower SA, with the exception of focal higher SA in bilateral precentral gyrus and insula in 22q11DS compared to controls (Figure 1B). The dispersion results also showed significantly greater within-group variance in SA in the left superior temporal, left transverse temporal, and right parahippocampal gyrus, but *lower* within-group variance in the bilateral caudal and left rostral anterior cingulate cortex. For regions with significant dispersion differences, the quantile shifts are visualized in Figures 2A-D for CT and Figures 3A-F for SA.

**Figure 2.**
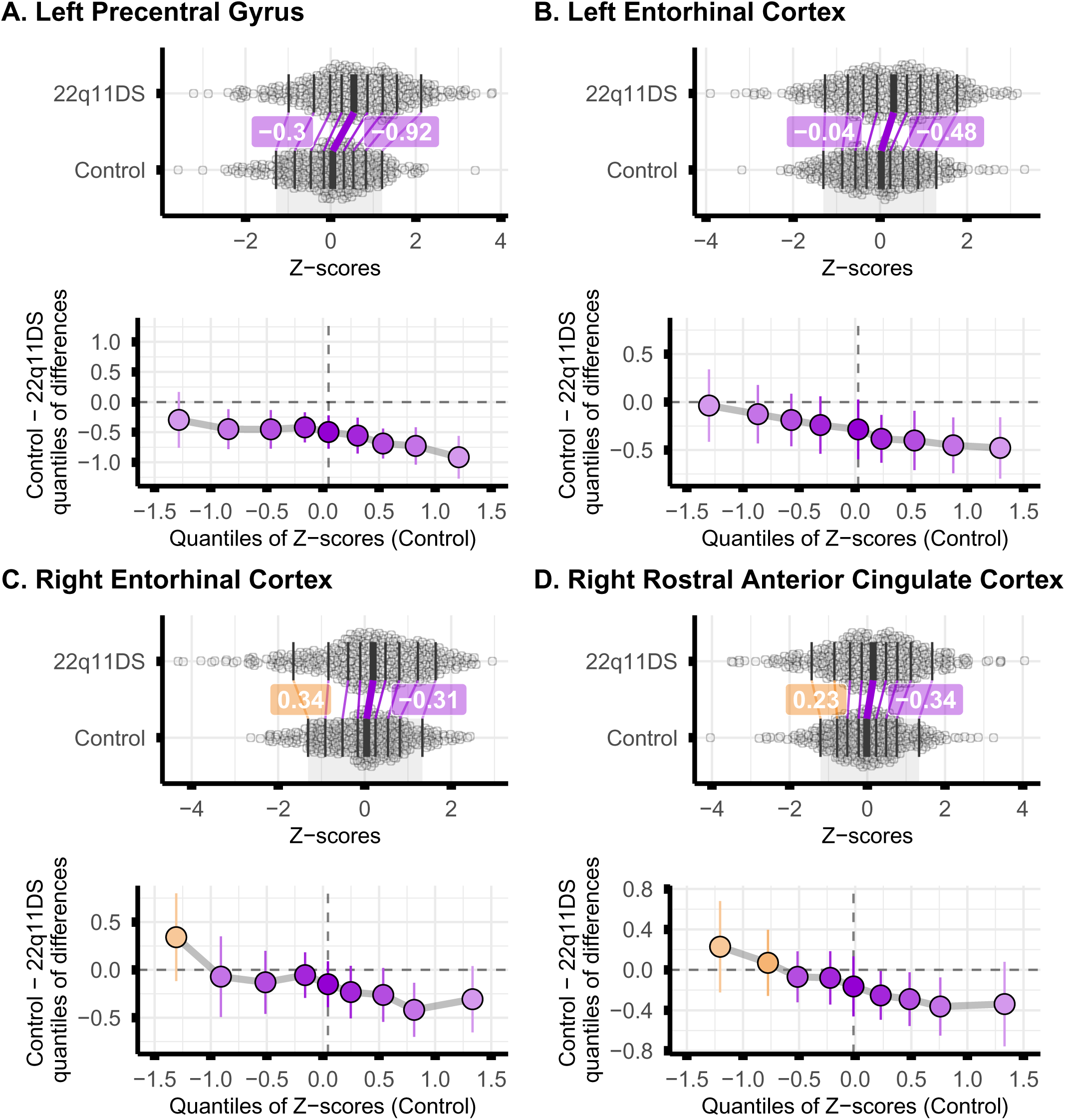
Quantile plots of regional cortical thickness. Quantile plots of the regions that showed significant dispersion differences in cortical thickness using double generalized linear models. Quantile plots of **A.** left precentral gyrus, **B.** left entorhinal cortex, **C.** right entorhinal cortex, and **D.** right rostral anterior cingulate, comparing 22q11DS to controls. Each vertical line represents the quantile difference from the 10th to 90th percentile in steps of 10, within each of the regions, showing how many units the 22q11DS group must be shifted to align with the corresponding quantiles of the control group (top figure). The vertical lines around the point estimates represent the 95% bootstrapped confidence intervals (bottom figure). The colors indicate the direction (orange = the quantile is lower in 22q11DS vs controls, purple = the quantile is greater in 22q11DS vs controls). The numeric labels (in highlighted boxes) represent the differences in the 10^th^ and 90^th^ quantiles, respectively. Here, quantile differences between 22q11DS and controls tend to grow larger at higher cortical thickness values.

**Figure 3.**
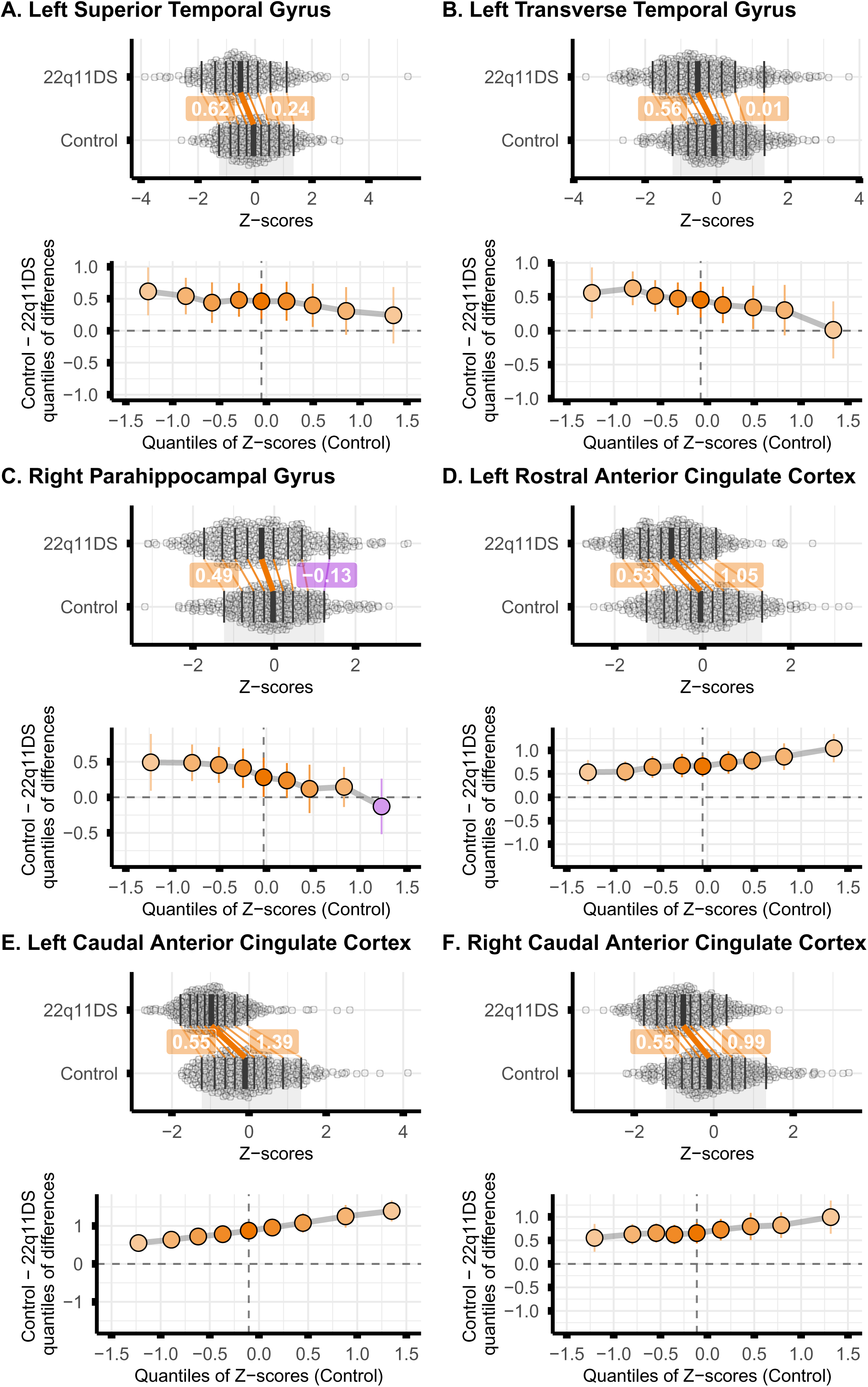
Quantile plots of regional cortical surface area. Quantile plots of the regions that showed significant dispersion differences in cortical surface area using double generalized linear models. Quantile plots of **A.** left superior temporal gyrus, **B.** left transverse temporal gyrus, **C.** right parahippocampal gyrus, and **D.** left rostral anterior cingulate, **E.** left caudal anterior cingulate, **F.** right caudal anterior cingulate, comparing 22q11DS to controls. Each vertical line represents the quantile difference from the 10th to 90th percentile in steps of 10, within each of the regions, showing how many units the 22q11DS group must be shifted to align with the corresponding quantiles of the control group (top figure). The vertical lines around the point estimates represent the 95% bootstrapped confidence intervals (bottom figure). The colors indicate the direction (orange = the quantile is lower in 22q11DS vs controls, purple = the quantile is greater in 22q11DS vs controls). The numeric labels (in highlighted boxes) represent the differences in the 10^th^ and 90^th^ quantiles, respectively. Here, the greatest quantile differences are observed in the lower quantiles for the left superior temporal gyrus, left transverse temporal gyrus and right parahippocampal gyrus (A-C), whereas the greatest quantile differences are observed in the upper quantiles for the three anterior cingulate subregions (D-F).

### Effects of disorder-related variables

We adjusted for diagnostic status (sFigures 3-4, eTable 3), current antipsychotic medication use (sFigures 5-6, eTable 4), and deletion subtype (sFigures 7-8, eTable 5) to assess whether these factors contributed to the heterogeneity in 22q11DS. Overall, the main findings for mean effects were largely unchanged. Mean differences in CT and SA remained significant across most regions after adjustment, with some attenuation in SA effects in frontal and inferior parietal regions, and in bilateral posterior cingulate and left transverse temporal CT. SA dispersion effects remained significant for left rostral and caudal anterior cingulate cortex and right transverse temporal after adjustment for diagnosis status, antipsychotic medication use, and deletion subtype, whereas right caudal anterior cingulate remained survived adjustment after antipsychotic medication use and deletion subtype, and right parahippocampal gyrus after deletion subtype. For CT dispersion, increased variability in the left precentral gyrus and right entorhinal cortex remained significant after adjustment for diagnostic status, antipsychotic medication use, and deletion subtype. Increased dispersion in the right rostral anterior cingulate cortex remained significant after adjustment for diagnostic status and medication use, but not deletion subtype.

We additionally examined the effects of ASD and psychosis diagnoses within the 22q11DS group. ASD diagnosis was not associated with significant differences in either mean CT or SA (sFigure 9-10, eTable 6-7). However, psychosis diagnosis (sFigures 11-12, eTable 8-9) and antipsychotic medication (sFigures 13-14, eTable 10-11) were associated with lower CT, with the strongest effect observed for psychosis diagnosis. Deletion subtype was not associated with mean differences in CT, but atypical deletion was associated with higher SA (sFigures 15-16, eTable 12-13). None of the clinical variables nor deletion subtype yielded significant dispersion effects.

### Spatial overlap between cortical maps

The cortical maps before and after the inclusion of additional covariates (e.g., psychiatric diagnosis) were highly correlated (*r* = 0.93 to 0.99, all *p*_spins_ < 0.001). For the remaining analyses, we used the cortical maps that were derived after adjusting for age, age^2^, sex, psychotic disorder, and ASD (+ intracranial volume for SA) to account for systematic regional mean shifts driven by neuropsychiatric diagnosis. There was no association between the mean CT map (regions that differed on average) and the dispersion CT map (regions with different variability around the mean, *r* = 0.16, *p*_spin_ = 0.128). However, the mean SA map was significantly associated with the dispersion SA map (*r* = 0.34, *p*_spin_ = 0.024), indicating that brain regions with larger SA tend to exhibit greater variability around the mean in 22q11DS relative to controls.

The mean CT map showed spatial correspondence with density maps of GABAergic (*r* = 0.39, *p*_spin_ = 0.003, Fig. 4A-B) and NMDA (*r* = 0.42, *p*_spin_ = 0.003, Fig 4C) receptors, as well as a suggestive correspondence with the density map of mGluR5 receptors (*r* = 0.24, *p*_spin_ = 0.066, Fig. 4D), indicating that thicker brain regions in 22q11DS tend to correspond to brain regions that are typically densely populated with receptors for the brain’s primary inhibitory and excitatory neurotransmitters. Thus, regions showing increased CT in 22q11DS vs controls overlap with regions that typically exhibit high excitatory-inhibitory synaptic density. The dispersion CT map showed spatial correspondence with the density map of GABAergic (*r* = –0.30, *p*_spin_ = 0.021, Fig. 4E), but not NMDA (*r* = –0.01, *p*_spin_ = 0.474) nor mGluR5 (*r* = 0.08, *p*_spin_ = 0.324) receptors. Thus, brain regions that exhibit greater CT variability in 22q11DS are depleted for receptors of GABAa. Since the secondary analyses revealed regional main effects of psychosis on CT, we also examined the spatial overlap between mean CT map for 22q11DS with psychosis and neurotransmitter density. Here, the mean effect of psychosis on CT spatially overlapped with NMDA (*r* = –.47, *p*_spin_ = .002, Fig. 4F) and mGluR5 (*r* = –.56, *p*_spin_ < .001, Fig. 4G), but not GABAa (*r* = –.08, *p*_spin_ = .33) receptor density, indicating that lower CT associated with psychosis in 22q11DS occurs in regions enriched for glutamatergic receptors.

**Figure 4.**
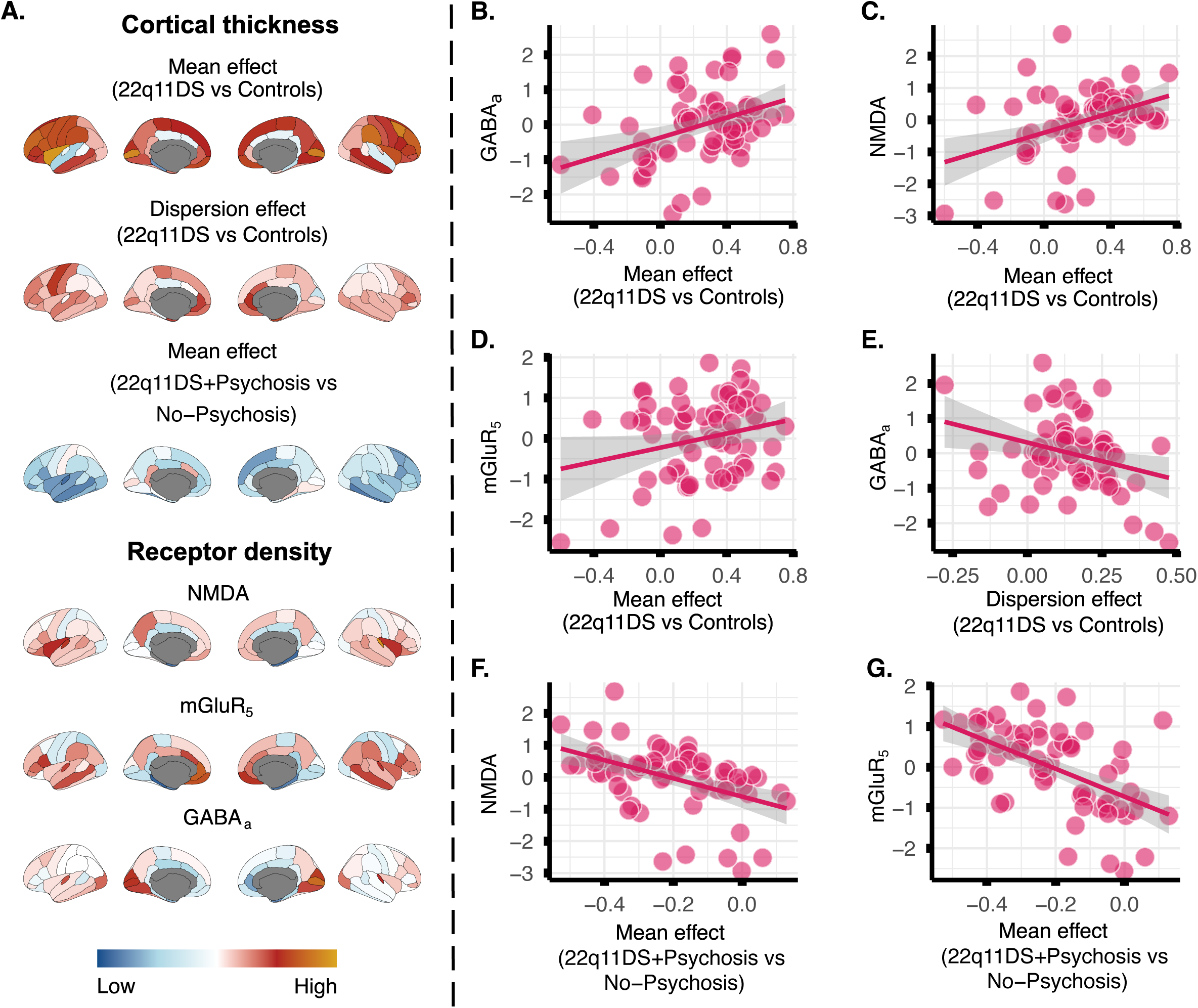
Spatial overlap between cortical maps. **A.** Overview of the cortical thickness and receptor density maps included in the correlation analyses. The cortical thickness maps reflect case-control differences (i.e., z-standardized differences), and receptor density maps reflect Z-scored receptor density values. Scatterplots showing that greater cortical thickness in 22q11DS vs controls align with regions exhibiting high **B.** GABAa, **C.** NMDA and **D.** mGluR5 (trend level) receptor density, whereas greater within-group variability in 22q11DS aligns with regions exhibiting lower **E.** GABAa receptor density. Regions that show lower cortical thickness in 22q11DS with psychosis vs no psychosis align with regions that are enriched for **F.** NMDA and **G.** mGluR5 receptors.

The mean SA map did not show significant spatial correspondence with any of the cortical expansion maps examined (*r* = .05 to .21, all *p*_spins_ > .05). While the dispersion SA map did not exhibit spatial correspondence with the developmental (*r* = .03, *p*_spin_ = .401) nor evolutionary (*r* = .07, *p*_spin_ = .311) expansion maps, it showed significant spatial correspondence with the NIH allometric scaling map (*r* = .31, *p*_spin_ = .013, sFigure 18A-B) and a suggestive but not statistically significant spatial correspondence with the PNC allometric scaling map (*r* = .27, *p*_spin_ = .055, sFigure 18C), indicating that regions that exhibit greater variance in SA among 22q11DS tend to correspond to regions that typically expand faster relative to overall brain size in humans.

## Discussion

In this large multisite study, we extend prior work in 22q11DS by demonstrating that the syndrome is characterized not only by systematic mean shifts in CT and SA (6), but also by region-specific alterations in the *distribution* of these measures. By applying double generalized linear models, we show that the neuroanatomical impact of 22q11DS is highly individualized for specific regions, possibly reflecting differential impact on cell-types that dominate these brain regions. At the same time, 22q11DS exerted a notably consistent effect on anterior cingulate SA, reflected by reduced dispersion relative to controls. Together, these findings suggest that the deletion produces both broadly shared developmental constraints and highly individualized effects on cortical maturation.

The most notable variability in CT emerged in precentral, entorhinal, and rostral anterior cingulate regions, suggesting that 22q11DS selectively destabilizes regional developmental programs. Importantly, dispersion effects on CT persisted after accounting for clinical covariates, suggesting that heightened regional variability is a core feature of the deletion, rather than a secondary consequence of illness or treatment. In contrast, SA variability in the left superior temporal gyrus, a region implicated in auditory processing and language(47,48), was consistently attenuated after covariate adjustment, indicating greater sensitivity to clinical or treatment-related factors. Previous studies of individuals with idiopathic schizophrenia have reported widespread brain structural heterogeneity beyond mean differences, which have been interpreted to reflect the heterogeneous etiologies of schizophrenia (9,10).

Not all effects reflected increased heterogeneity. Reduced dispersion of anterior cingulate SA suggests that 22q11DS tightly constrains development of this region, making affected individuals more neuroanatomically similar than controls. Interestingly, reduced variability in cingulate morphology (folding) has also been reported in psychosis-spectrum disorders compared to controls (10). As folding and SA share a common developmental origin (49), altered cingulate development may represent a shared neurobiological pathway related to psychosis risk.

CT differences placed in a neurochemical context revealed a structured pattern. The spatial correspondence between brain maps show that the main and dispersion effects of 22q11DS on brain structure aligns with excitatory-inhibitory neurotransmitter density, possibly pointing towards a role of glutamate and GABAergic neurotransmitter circuits in the cortical alterations in 22q11DS(43–46). Regions showing cortical thickening were preferentially localized to brain regions rich in NMDA and GABAergic receptors, whereas regions exhibiting the greatest inter-individual variability were relatively depleted of GABAergic receptors. This dissociation suggests that the impact of 22q11DS on cortical maturation is spatially patterned by the underlying excitatory–inhibitory (E/I) architecture. Our analysis also complements previous findings that have shown a high contribution of serotonin neurotransmitter systems to the spatial map of 22q11DS using dominance analysis (42), as well as striatal dopaminergic alterations(50). Notably, we found that cortical thinning associated with psychosis in 22q11DS is localized to glutamatergic-dense cortical regions, consistent with models of exaggerated synaptic remodeling in excitatory networks in idiopathic schizophrenia. These findings align with MR spectroscopy studies showing altered levels of glutamate+glutamine in idiopathic schizophrenia, first-episode psychosis (51,52), ASD (53,54) and 22q11DS (55) compared to controls, as well as alterations in GABA-ergic inhibition measured using TMS-EMG and functional indices of E/I balance derived from rs-fMRI in patients with psychosis (56) and ASD (57,58). Together, these findings support the relevance of E/I-related mechanisms, although the present analyses do not establish causality.

Lastly, by aligning SA dispersion with cortical scaling maps, we find partial support for spatial correspondence between SA dispersion and allometric scaling maps. As SA is largely established by early childhood (59), case-control differences in SA measures should be interpreted within the context of early cortical development, where association cortices show greater evolutionary and developmental expansion compared to sensorimotor regions (32). As allometric scaling reflects regional expansion or constraint with increasing brain size, our results suggest that 22q11DS amplifies the constraints on regions that already show slow expansion rate in the population instead of affecting regions that have expanded the most during human evolution or show prolonged developmental maturation. This could indicate that 22q11DS has particularly strong regional-specific effects, such as on the cingulate, possibly due to a disruption to the radial glia pool influencing the expansion of the human cortex (12,13,60). Interestingly, the insular cortex, which is also found to develop early and exhibit hypoallometric scaling (61), is larger in 22q11DS vs controls after adjusting for intracranial volume. While speculative, this pattern implicates early perturbations in radial glial–mediated tangential growth and suggests that genetically driven constraints interact with region-specific progenitor programs to shape the cortical phenotype of 22q11DS.

## Limitations

Several limitations should be considered. Spatial correspondence analyses relied on independently derived receptor and developmental atlases and therefore cannot establish causal relationships. Structural variability may also reflect unmeasured environmental, developmental, or methodological factors. However, we minimized these effects through harmonized preprocessing, quality control, and site correction procedures, and the principal findings remained robust after adjustment for major clinical covariates.

## Conclusions

Together, our findings establish that 22q11DS sculpts the cortex through a dual mechanism of mean shifts and altered variability, patterned by neurochemical and developmental architectures. Ultimately, recognizing heterogeneity in brain structure is essential for unraveling the complex neurobiological pathways from genotype to neuropsychiatric phenotype in 22q11DS. Cortical regions showing reduced variability may reflect core neurobiological features of the syndrome, whereas heightened variability may index processes contributing to the marked heterogeneity of neuropsychiatric outcomes in this population.

## Data Availability

Data were derived from the ENIGMA-22q Working Group from cohorts contributing structural MRI data. Requests for individual level data must be directed to the individual site PIs.

## Acknowledgements

This project was supported by the Stephen R. Mallory Award for Schizophrenia Research (RB), National Institute of Mental Health (Grant Nos. R01/R37MH085953, R01MH129858, U01MH101779, U01MH119736, and R21MH116473, CEB), the Simons Foundation Autism Research Initiative Explorer Award (CEB), Uytengsu-Hamilton 22q11 Neuropsychiatry Research Award (CEB), F31DA060068 (CMA), Eunice Kennedy Shriver National Institute of Child Health & Human Development, RO1HD42974 (TJS), Canadian Institutes of Health Research (CIHR) grants MOP-79518, MOP-89066, MOP-97800 and MOP-111238 (ASB), the Medical Wellcome Trust Discovery Award 227882/Z/23/Z (MBMvdB, MJO), Wellcome Trust 226709/Z/22/Z(MBMvdB, MJO), MRC MR/W028395/1 (MBMvdB, MJO), MRC MR/W020297/1(MBMvdB, MJO), MRC MR/W014416/1(MBMvdB, MJO), MRC MR/T033045/1(to MBMvdB, MJO, SJRAC), MRC MR/S037667/1(MBMvdB, MJO), NIMH U01MH119758 (MBMvdB, MJO). National Institutes of Health (R21 MH139001, R01 MH129742, R01 MH131806, R01 AG058854, R01 MH134962, CRKC), the Office Of The Director, National Institutes Of Health of the National Institutes of Health under Award Number S10OD032285 (CRKC), FONDECYT regular 1240426 from Agencia Nacional de Investigación y Desarrollo (ANID) Chile (NAC), and the Centro de Interés Nacional grant IINARA, CIN250068, ANID Chile (NAC), R01 MH064824 (WRK), R01MH085953 (LK), R01-MH134965-01 (DMM-M); RO1-MH119185-01A1 (DMM-M); U01-MH119737-01 (DMM-M), ANID-Chile Fondecyt grants 1171014 and 1211014 (GMR), NIMH 5R01MH133843 (JES), R01MH107018 (TJS), R01MH129858 (PMT), NSF Graduate Research Fellowship (ZHT), R01MH100900 (JFH), NIH GM125757 (BSE), The National Institute of Mental Health: International Consortium on Brain and Behavior in 22q11.2 Deletion Syndrome 5U01MH101722 (JV), NIH Training Fellowship 2T32AG058507-06 (JEVR).

## ECHO-DEFINE

This work was supported by the Wellcome Trust (Institutional Strategic Support Fund [to MBMvdB] and Clinical Research Training Fellowship Grant No. 102003/Z/13/Z [to JLD]), Waterloo Foundation (Grant No. WF 918-1234 [to MBMvdB]), Baily Thomas Charitable Fund (Grant No. 2315/1 [to MBMvdB]), NIMH (Grant Nos. 5UO1MH101724 and U01MH119738 [to MBMvdB]), IMAGINE-ID and IMAGINE-2 studies (funded by Medical Research Council Grant Nos. MR/N022572/1 and MR/T033045/1 [to MBMvdB]), and Medical Research Council (Centre Grant No. MR/P005748/1 [to MJO]). The DEFINE study was supported by a Wellcome Trust Strategic Award (Grant No. 100202/Z/12/Z [to MJO])

